# Longitudinal characteristics of lymphocyte responses and cytokine profiles in the peripheral blood of SARS-CoV-2 infected patients

**DOI:** 10.1101/2020.02.16.20023671

**Authors:** Jing Liu, Sumeng Li, Jia Liu, Boyun Liang, Xiaobei Wang, Hua Wang, Wei Li, Qiaoxia Tong, Jianhua Yi, Lei Zhao, Lijuan Xiong, Chunxia Guo, Jin Tian, Jinzhuo Luo, Jinghong Yao, Ran Pang, Hui Shen, Cheng Peng, Ting Liu, Qian Zhang, Jun Wu, Ling Xu, Sihong Lu, Baoju Wang, Zhihong Weng, Chunrong Han, Huabing Zhu, Ruxia Zhou, Helong Zhou, Xiliu Chen, Pian Ye, Bin Zhu, Shengsong He, Yongwen He, Shenghua Jie, Ping Wei, Jianao Zhang, Yinping Lu, Weixian Wang, Li Zhang, Ling Li, Fengqin Zhou, Jun Wang, Ulf Dittmer, Mengji Lu, Yu Hu, Dongliang Yang, Xin Zheng

**Author notes:** These authors contribute equally to this work (Jing Liu, Sumeng Li, Jia Liu, Yu Hu, Dongliang Yang, Xin Zheng). **Correspondence to:** Prof. Dr. Yu Hu, Department of Hematology, Union Hospital, Tongji Medical College, Huazhong University of Science and Technology, Wuhan 430022, China, Prof. Dr. Dongliang Yang, Department of Infectious Diseases, Union Hospital, Tongji Medical College, Huazhong University of Science and Technology, Wuhan 430022, China, Prof. Dr. Xin Zheng, Department of Infectious Diseases, Union Hospital, Tongji Medical College, Huazhong University of Science and Technology, Wuhan 430022, China.

## Abstract

**Background:** The dynamic changes of lymphocyte subsets and cytokines profiles of patients with novel coronavirus disease (COVID-19) and their correlation with the disease severity remain unclear.

**Methods:** Peripheral blood samples were longitudinally collected from 40 confirmed COVID-19 patients and examined for lymphocyte subsets by flow cytometry and cytokine profiles by specific immunoassays.

**Results:** Of the 40 COVID-19 patients enrolled, 13 severe cases showed significant and sustained decreases in lymphocyte counts but increases in neutrophil counts than 27 mild cases. Further analysis demonstrated significant decreases in the counts of T cells, especially CD8 + T cells, as well as increases in IL-6, IL-10, IL-2 and IFN-γ levels in the peripheral blood in the severe cases compared to those in the mild cases. T cell counts and cytokine levels in severe COVID-19 patients who survived the disease gradually recovered at later time points to levels that were comparable to those of the mild cases. Moreover, the neutrophil-to-CD8+ T cell ratio (N8R) were identified as the most powerful prognostic factor affecting the prognosis for severe COVID-19.

**Conclusions:** The degree of lymphopenia and a proinflammatory cytokine storm is higher in severe COVID-19 patients than in mild cases, and is associated with the disease severity. N8R may serve as a useful prognostic factor for early identification of severe COVID-19 cases.

**Summary:** Lymphocyte subsets and cytokine profiles in the peripheral blood of COVID-19 patients were longitudinally characterized. The study revealed the kinetics features of immune parameters associated with the disease severity and identified N8R as a useful prognostic factor for predicting severe COVID-19 cases.

## Introduction

First reported in Wuhan, China, on 31 December 2019, an ongoing outbreak of a viral pneumonia in humans has raised acute and grave global concern. The causative pathogen was rapidly identified as a novel β-coronavirus, which has since been formally named as the severe acute respiratory syndrome coronavirus 2 (SARS-CoV-2) by the International Committee on Taxonomy of Viruses. According to the daily report of the National Health Commission of China, the epidemic of SARS-CoV-2 has so far caused 57,416 confirmed cases, including 11,272 severe cases, and 1,665 deaths in China by February 15th, 2020[1]. The disease caused by SARS-CoV-2 has been recently named as the Coronavirus Disease-2019 (COVID-19) by the World Health Organization. Previous studies about the epidemiological and clinical characteristics of COVID-19 showed patients with COVID-19 may develop either mild or severe symptoms of acute respiratory infection, while the mild patients show symptoms of fever, dry cough, fatigue, abnormal chest CT findings but with a good prognosis[2,3]. In contrast, some patients develop severe pneumonia, acute respiratory distress syndrome (ARDS) or multiple organ failure, with death rates ranging from between 4.3% to 15% according to different study reports[2,4].

Lymphopenia and inflammatory cytokine storm are typical laboratory abnormalities observed during highly pathogenic coronavirus infections, such as the severe acute respiratory syndrome coronavirus (SARS-CoV) and the Middle East respiratory syndrome coronavirus (MERS-CoV) infections, and are believed to be associated with disease severities[5,6]. Recent studies have also reported decreases in the counts of lymphocytes in the peripheral blood and increases in serum inflammatory cytokine levels in COVID-19 patients[4,7]. However, it has remained largely unclear how different lymphocyte subsets and the kinetics of inflammatory cytokines change in the peripheral blood during COVID-19. In this study, we longitudinally characterized the changes of lymphocyte subsets and cytokines profiles in the peripheral blood of COVID-19 patients with distinct disease severities.

## Methods

### Data collection

A written informed consent was regularly obtained from all patients upon admission into Wuhan Union Hospital, China. The study was approved by the Ethics Committee of Tongji Medical College of Huazhong University of Science and Technology. The 40 confirmed COVID-19 patients at Wuhan Union Hospital during January 5 to January 24, 2020 were enrolled into this retrospective single-center study. All medical record information including epidemiological, demographic, clinical manifestation, laboratory data, and outcome data were obtained. All data were checked by a team of trained physicians.

### Laboratory examination

Laboratory confirmation of the SARS-CoV-2 was performed by local CDC according to Chinese CDC protocol. Throat-swab specimens were collected from all patients and the samples were maintained in viral-transport medium for laboratory testing. An infection with other respiratory viruses including influenza A virus, influenza B virus, Coxsackie virus, respiratory syncytial virus, parainfluenza virus and enterovirus was excluded by real-time RT-PCR. Specimens, including sputum or alveolar lavatory fluid, blood, urine, and feces, were cultured to identify pathogenic bacteria or fungi that may be associated with the SARS-CoV-2 infection. The specific IgG and IgM of Chlamydia pneumonia and Mycoplasma pneumonia were detected by chemiluminescence immunoassay. The lymphocyte test kit (Beckman Coulter Inc., FL, USA) was used for lymphocyte subset analysis. Plasma cytokines (IL2, IL4, IL6, IL10, TNF - α and IFN - γ) were detected with human Th1/2 cytokine kit II (BD Ltd., Franklin lakes, NJ, USA). All tests are performed according to the product manual.

### Statistical analyses

Classification variables are expressed in frequency or percentage, and significance is detected by chi square or Fisher’s exact test. The quantized variables of parameters are expressed as mean ± standard deviation, and the significance is tested by t-test. Nonparametric variables were expressed in median and quartile intervals, and significance was tested by Mann Whitney U or Kruskal Wallis test. Data (nonnormal distribution) from repeated measures were compared using the generalized linear mixed model. P < 0.05 was considered statistically significant in all statistical analyses. Principal component analysis (PCA) was performed to identify the major contributing factors among clinical parameters to distinguish mild and severe cases of COVID-19 patients. The diagnostic values of selected parameters for differentiating mild and severe cases of COVID-19 patients were assessed by receiver operating characteristic (ROC) and area under the ROC curve (AUC). SPSS statistical software (Macintosh version 26.0, IBM, Armonk, NY, USA) and R package are used for statistical analysis.

## Results

### Demographic and clinical characteristics of COVID-19 patients

The diagnosis of COVID-19 for patients was performed according to the Guidelines of the Diagnosis and Treatment of New Coronavirus Pneumonia (version 5) published by the National Health Commission of China. Mild patients met all following conditions: (1) Epidemiology history, (2) Fever or other respiratory symptoms, (3) Typical CT image abnormities of viral pneumonia, and (4) Positive result of RT-PCR for SARS-CoV-2 RNA. Severe patients additionally met at least one of the following conditions: (1) Shortness of breath, RR≥30 times/min, (2) Oxygen saturation (Resting state) ≤93%, or (3) PaO2 / FiO2 ≤300mmHg. A total of 40 patients were enrolled in this study, which were all Wuhan residents and laboratory confirmed cases. The patients were divided into two groups according to above-mentioned conditions, including 27 mild cases (67.5%) and 13 severe cases (32.5%). Two patients in the severe group died on day 15 and 21 after disease onset.

The enrolled COVID-19 patients consisted of 15 males (37.5%) and 25 females (62.5%) (Table 1). Only 3 patients (7.5%) had an exposure history (shopping) on the Huanan seafood market in Wuhan. The medium age of the patients was 48.7 ± 13.9 years old. The ages of the severe patient group (59.7 ± 10.1 years) were older than that of the mild group (43.2 ± 12.3 years). A total of 14 (35%) patients in both groups had basic diseases, including diabetes (6 [15%]), hypertension (6 [15%]), pituitary adenoma (2 [5%]), thyroid disease (2 [5%]) and tumor disease (2 [5%]). Four severe patients had mixed fungal infection and 1 severe patient had mixed bacterial infection (Table 1). All severe patients and 85.2% of the mild patients had fever, while no significant difference in the degrees of temperature was observed between the two groups (Table 1). The severe patients showed significantly higher frequencies in the occurrence of sputum production (p=0.032), myalgia (p=0.041) and nausea (p=0.029) (Table 1). The levels of fibrinogen (p<0.001), D-dimer (P=0.008), total bilirubin (p=0.007), aspartate transaminase (p<0.001), alanine transaminase (p=0.004), lactate dehydrogenase (p<0.001), creatine kinase (p=0.010), C-reactive protein (p=0.006), ferritin (p=0.015) and serum amyloid A protein (SAA, p=0.003) in the peripheral blood of the severe patients were significantly higher at admission compared to the mild patients (Table 2). No significant differences in the serum levels of immunoglobulins (IgA, IgG and IgM), complement C3 or C4 were observed between the two groups (Table 2).

**Table 1.** Demographics and baseline characteristics of patients infected with SARS-CoV-2.

| Baseline variables | All patients<br>(N=40) | Mild patients<br>(N=27) | Severe patients<br>(N=13) | P-value |
| --- | --- | --- | --- | --- |
| Characteristics |  |  |  |  |
| Age (year) | 48.7 ± 13.9 | 43.2 ± 12.3 | 59.7 ± 10.1 | <0.001 |
| Gender (%) |  |  |  | 0.138 |
| Men | 15 (37.5) | 8 (29.6) | 7 (53.8) |  |
| Women | 25 (62.5) | 19 (70.4) | 6 (46.2) |  |
| Huanan seafood market exposure (%) | 3 (7.5) | 1 (3.7) | 2 (5.4) | 0.242 |
| Underlying diseases (%) | 14 (35.0) | 7 (25.9) | 7 (53.8) | 0.155 |
| Diabetes | 6 (15.0) | 2 (7.4) | 4 (30.8) | 0.075 |
| Hypertension | 6 (15.0) | 1 (3.7) | 5 (38.5) | 0.010 |
| Pituitary adenoma | 2 (5.0) | 1 (3.7) | 1 (7.7) | >0.999 |
| Thyroid disease | 2 (5.0) | 2 (7.4) | 0 | >0.999 |
| Malignancy | 2 (5.0) | 2 (7.4) | 0 | >0.999 |
| Co-infection (%) | 5 (12.5) | 0 | 5 (38.5) | 0.002 |
| Fungi | 4 (10.0) | 0 | 4 (30.8) | 0.008 |
| Bacteria | 1 (2.5) | 0 | 1 (7.7) | >0.999 |
| Signs and symptoms |  |  |  |  |
| Fever | 36 ( 90.0 ) | 23 ( 85.2 ) | 13 ( 100 ) | 0.284 |
| Highest temperature, °C |  |  |  |  |
| <37.3 | 4 ( 10.0 ) | 4 ( 14.8 ) | 0 | 0.284 |
| 37.3–38.0 | 10 ( 25 ) | 8 ( 29.6 ) | 2 ( 15.4 ) | 0.451 |
| 38.1–39.0 | 17 ( 42.5 ) | 9 ( 33.3 ) | 8 ( 61.5 ) | 0.091 |
| >39.0 | 9 ( 22.5 ) | 6 ( 22.2 ) | 3 ( 23.1 ) | >0.999 |
| Chill | 10 ( 25 ) | 5 ( 18.5 ) | 5 ( 38.5 ) | 0.246 |
| Shivering | 5 ( 12.5 ) | 2 ( 7.4 ) | 3 ( 23.1 ) | 0.307 |
| Fatigue | 22 ( 55 ) | 14 ( 51.9 ) | 8 ( 61.5 ) | 0.564 |
| Cough | 33 ( 82.5 ) | 22 ( 81.5 ) | 11 ( 84.6 ) | >0.999 |
| Sputum production | 21 ( 52.5 ) | 11 ( 40.7 ) | 10 ( 76.9 ) | 0.032 |
| Pharyngalgia | 5 ( 12.5 ) | 4 ( 14.8 ) | 1 ( 7.7 ) | >0.999 |
| Dizziness | 7 ( 17.5 ) | 4 ( 14.8 ) | 3 ( 23.1 ) | 0.662 |
| Headache | 8 ( 20.0 ) | 6 ( 22.2 ) | 2 ( 15.4 ) | >0.999 |
| Rhinorrhea | 1 ( 2.5 ) | 1 ( 3.7 ) | 0 | >0.999 |
| Chest tightness | 12 ( 30.0 ) | 7 ( 25.9 ) | 5 ( 38.5 ) | 0.476 |
| Chest pain | 1 ( 2.5 ) | 1 ( 3.7 ) | 0 | >0.999 |
| Shortness of breath | 5 ( 12.5 ) | 5 ( 18.5 ) | 0 | 0.154 |
| Dyspnoea | 1 ( 2.5 ) | 1 ( 3.7 ) | 0 | >0.999 |
| Myalgia | 15 ( 37.5 ) | 7 ( 25.9 ) | 8 ( 61.5 ) | 0.041 |
| Abdominal pain | 1 ( 2.5 ) | 1 ( 3.7 ) | 0 | >0.999 |
| Diarrhoea | 3 ( 7.5 ) | 1 ( 3.7 ) | 2 ( 15.4 ) | 0.242 |
| Nausea | 3 ( 7.5 ) | 0 | 3 ( 23.1 ) | 0.029 |
| Vomiting | 1 ( 2.5 ) | 0 | 1 ( 7.7 ) | 0.325 |
| Hypoleucocytosis | 10 ( 25.0 ) | 8 ( 29.6 ) | 2 ( 15.4 ) | 0.451 |
| Lymphopenia | 21 ( 52.5 ) | 11 ( 40.7 ) | 10 ( 76.9 ) | 0.046 |
| Thrombocytopenia | 5 ( 12.5 ) | 3 ( 11.1 ) | 2 ( 15.4 ) | >0.999 |

**Table 2.** Comparison of laboratory parameters between mild and severe COVID-19 patients.

| Baseline variables | All patients<br>(N=40) | Mild patients<br>(N=27) | Severe patients<br>(N=13) | P-value |
| --- | --- | --- | --- | --- |
| Hemoglobin (g/l) | 126.4 ± 13.4 | 127.8 ± 13.1 | 123.4 ± 14.0 | 0.334 |
| Platelet (×10<sup>9</sup>/L) | 183.1 ± 69.0 | 181.4 ± 70.7 | 186.6 ± 68.1 | 0.826 |
| White blood cell<br>(×10<sup>9</sup>/L) | 4.8 ± 2.6 | 3.9 ± 1.5 | 6.6 ± 3.4 | 0.002 |
| Neutrophil (×10<sup>9</sup>/L) | 2.8 (1.6-4.3) | 2.0 (1.5-2.9) | 4.7 (3.6-5.8) | <0.001 |
| Lymphocyte (×10<sup>9</sup>/L) | 0.9 (0.7-1.3) | 1.1 (0.8-1.4) | 0.6 (0.6-0.8) | 0.002 |
| Monocyte (×10<sup>9</sup>/L) | 0.3 (0.2-0.5) | 0.3 (0.2-0.5) | 0.2 (0.2-0.5) | 0.477 |
| TBil (umol/l) | 10.3 ± 5.0 | 8.8 ± 4.1 | 13.2 ± 5.5 | 0.007 |
| ALT (U/L) | 22.5 (16.8-31.2) | 19.0 (13.5-26.0) | 27.0 (23.0-50.0) | 0.004 |
| AST (U/L) | 34.1 ± 17.7 | 25.9 ± 9.5 | 51.2 ± 18.7 | <0.001 |
| LDH (U/L) | 303.9 ± 168.8 | 221.5 ± 71.2 | 462.4 ± 190.6 | <0.001 |
| CK (U/L) | 59.5 (45.0-88.8) | 51.0 (45.0-68.0) | 104.0<br>(77.0-124.0) | 0.010 |
| Blood urea nitrogen<br>(mmol/l) | 3.2 (2.5-4.3) | 3.2 (2.5-4.4) | 3.3 (2.7-3.7) | 0.707 |
| Serum creatinine (umol/l) | 67.3 ± 19.7 | 64.0 ± 13.3 | 74.2 ± 28.3 | 0.128 |
| Blood potassium (mmol/l) | 3.8 ± 0.5 | 3.9 ± 0.5 | 3.7 ± 0.4 | 0.242 |
| Blood sodium (mmol/l) | 145.9 ± 43.4 | 149.5 ± 52.5 | 138.6 ± 6.2 | 0.462 |
| D-Dimer (mg/l) | 0.6 (0.3-0.9) | 0.4 (0.2-0.8) | 0.9 (0.7-1.5) | 0.008 |
| PT (s) | 13.2 ± 0.6 | 13.1 ± 0.6 | 13.4 ± 0.6 | 0.154 |
| APTT (s) | 39.5 ± 4.5 | 39.5 ± 4.6 | 39.5 ± 4.2 | 0.968 |
| INR | 1.0 ± 0.1 | 1.0 ± 0.1 | 1.0 ± 0.1 | 0.154 |
| FIB (g/l) | 5.1 ± 1.6 | 4.5 ± 1.4 | 6.3 ± 1.3 | <0.001 |
| IgE | 43.9 (14.4-98.0) | 26.5 (12.8-76.1) | 43.9<br>(27.0-105.5) | 0.243 |
| IgG | 11.1 ± 2.0 | 10.8 ± 2.0 | 11.5 ± 2.0 | 0.370 |
| IgA | 2.2 ± 0.7 | 2.2 ± 0.8 | 2.4 ± 0.6 | 0.483 |
| IgM | 1.1 ± 0.4 | 1.1 ± 0.5 | 1.1 ± 0.3 | 0.918 |
| C-reactive protein (mg/l) | 38.1 (4.7-65.2) | 7.6 (3.1-57.3) | 62.9 (42.4-86.6) | 0.006 |
| Ferritin (ug/l) | 596.5<br>(308.6-1087.6) | 367.8<br>(174.7-522.0) | 835.5<br>(635.4-1538.8) | 0.015 |
| SAA (mg/l) | 134.4<br>(35.7-586.3) | 46.9<br>(20.5-134.4) | 607.1<br>(381.9-686.2) | 0.003 |
| C3 (g/l) | 0.8 ± 0.2 | 0.8 ± 0.2 | 0.8 ± 0.1 | 0.389 |
| C4 (g/l) | 0.3 ± 0.1 | 0.3 ± 0.1 | 0.3 ± 0.1 | 0.426 |

### Kinetic analysis of lymphocyte subsets in the peripheral blood of COVID-19 patients

Lymphopenia was observed in 44.4% (12/27) of mild patients and 84.6% (11/13) of severe patients at the onset of the disease. As shown in Table 2, the absolute counts of lymphocytes in the peripheral blood of the severe patients was significantly lower, while the absolute counts of total white blood cells (WBCs) and neutrophils were significantly higher, than those of the mild patients at the time of hospital admission. No significant difference in monocyte counts was observed between the two groups (Table 2). Next, we analyzed the kinetic changes of WBCs, neutrophils and monocytes as well as different lymphocyte subsets in the peripheral blood of COVID-19 patients from the disease onset to at least 16 days later. The two mortalities in the severe group were excluded from the analysis due to the lack of kinetic data. Significant increases in total WBCs counts in the severe group were only observed at the time point of onset (within 3 days) but not during the following period of disease progression compared to the mild group (Figure 1A). Significant increases in neutrophil counts of the severe group were observed not only at the time point of disease onset, but also at 13-15 days after compared to the mild group (Figure 1B). In contrast, a sustained decrease in lymphocyte counts of the severe group was observed compared to those of the mild patients. The difference was significant at the time point of disease onset and became even greater on 4-6 days later (Figure 1C). From 7-15 days after disease onset, the lymphocyte counts gradually increased in the severe group, and reached a comparable level to that of the mild patients at 16 days after disease onset (Figure 1C). No significant differences in monocyte counts were observed between the two groups during the whole observation period (Figure 1D).

**Figure 1.**
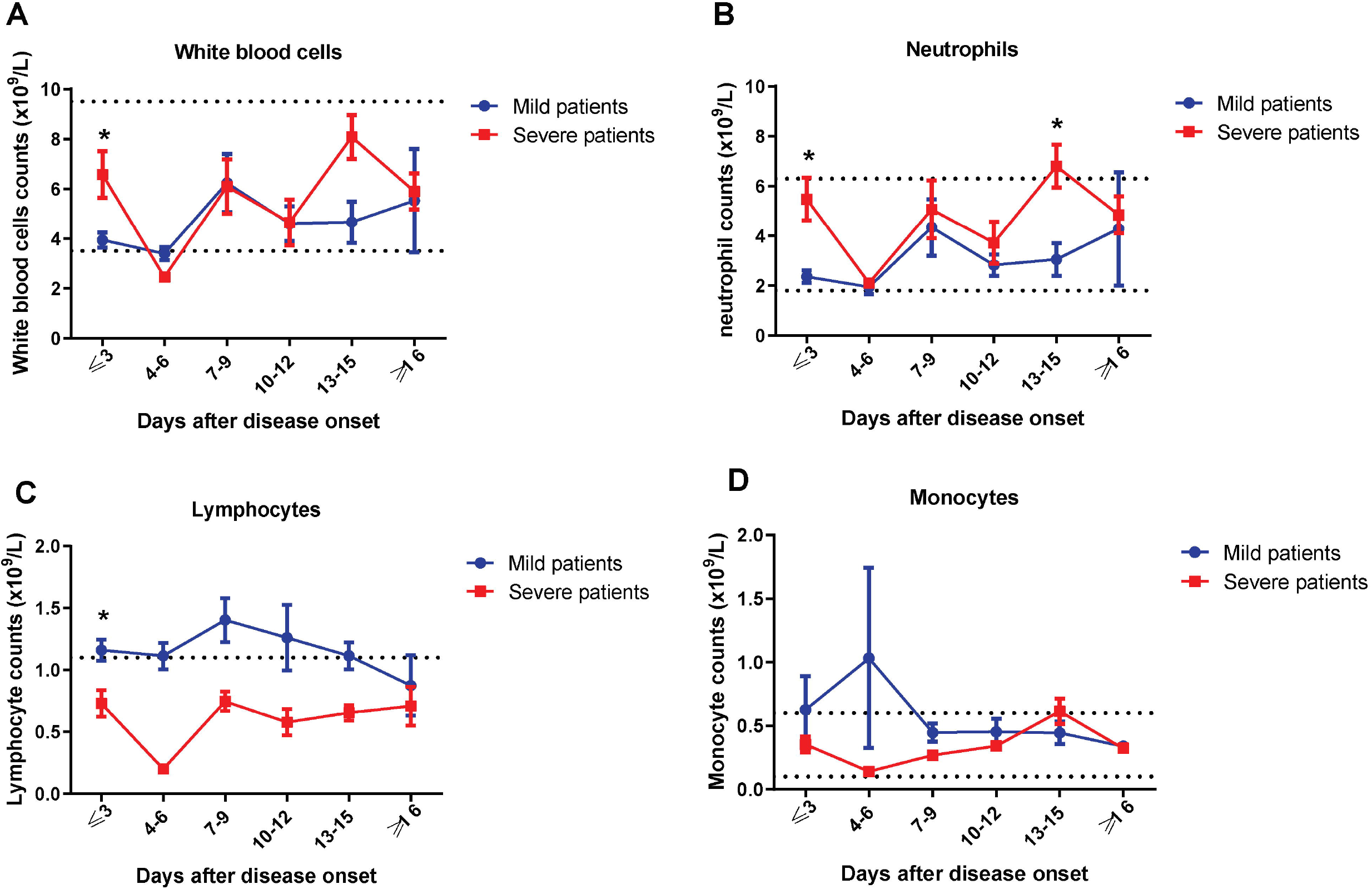
Kinetic analysis of cell counts of different populations of WBCs in COVID-19 patients. The absolute numbers of total WBCs (A), neutrophils (B), lymphocytes (C) and monocytes (D) in the peripheral blood of mild (blue line) and severe (red line) COVID-19 patients were analyzed at different time points after hospital admission. Error bars, mean ± s.e.m.; *p<0.05. The upper dotted lines show the upper normal limit of each parameter, and the lower dotted lines show the lower normal limit of each parameter.

In order to further determine the kinetic changes of different lymphocyte subsets in the peripheral blood of COVID-19 patients, we performed flow cytometry to stain CD3^+^ T cells, CD4^+^ and CD8^+^ T cell subsets, B cells and NK cells. Similar to the findings for lymphocytes, sustained decreases in CD3, CD8 and CD4 T cell counts was observed in the severe group compared to those of the mild patients during clinical observation (Figure 2A-C, Supplementary figure 1). The lowest CD3, CD4 and CD8 T cell counts were observed at 4-6 days after disease onset (Figure 2A-C). The differences in CD3 and CD8 T cell counts between the two groups were significant at the time points of disease onset and 7-9 days later (Figure 2A and 2C). However, the differences in CD4 T cell counts between the two groups did not reach a statistical significance at any time point (Figure 2C). The T cell counts started to gradually increase in the severe group starting at 7 days after disease onset, and reached comparable levels to those in the mild patients on day 16 after disease onset (Figure 2A-C). No significant differences in B cell and NK cell counts were observed between the two groups during the whole course of the disease (Figure 2D and 2E).

**Figure 2.**
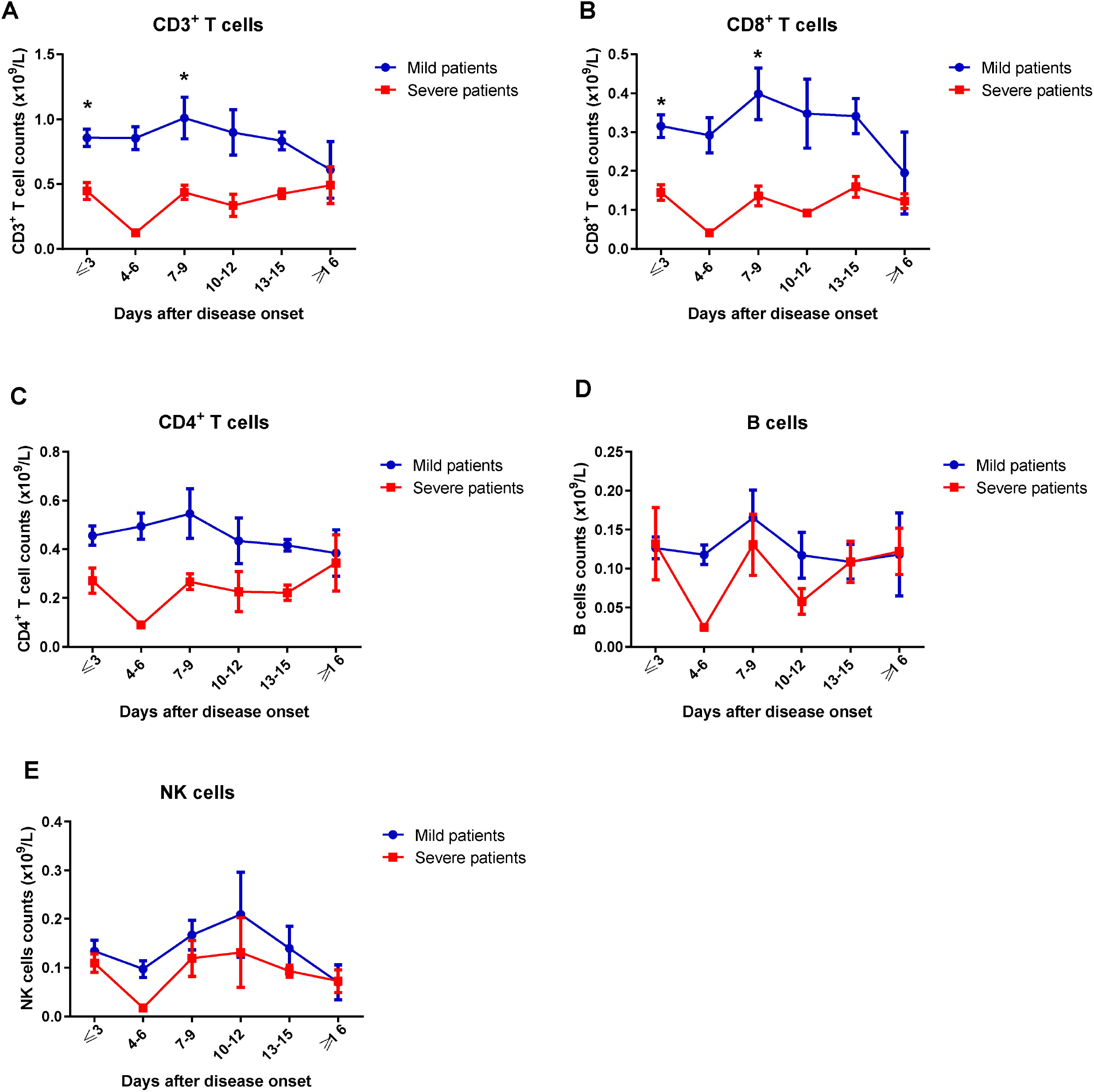
Kinetic analysis of cell counts of different lymphocyte subsets in COVID-19 patients. The absolute numbers of CD3+ T cells (A), CD8+ T cells (B), CD4+ T cells (C), B cells (D) and NK cells (E) in the peripheral blood of mild (blue line) and severe (red line) COVID-19 patients were analyzed at different time points after hospital admission. Error bars, mean ± s.e.m.; *p<0.05.

### Kinetic analysis of inflammatory cytokine levels in the serum of COVID-19 patients

A previous study demonstrated changes in inflammatory cytokine levels, such as IL-2, IL-7, IL-10, and TNF-α, in the serum of COVID-19 patients[2]. Therefore, we further characterized the kinetic changes of inflammatory cytokine levels, including IL-2, IL-4, IL-6, IL-10, IFN-γ and TNF-α, in the serum of our patient cohort. Fluctuations in the serum levels of these cytokines in the mild patient group were minor. In contrast, the severe patient group showed more significant fluctuations in the serum levels of these cytokines (Figure 3). All examined cytokines, except IL-6, reached their peak levels in the serum at 3-6 days after disease onset (Figure 3). Both IL-6 and IL-10 levels showed sustained increases in the severe group compared to the mild group (Figure 3A and 3B). A decease in serum IL-6 levels in the severe group started at 16 days after disease onset, and IL-10 levels were lowest at 13 days after disease onset (Figure 3A and 3B). Significant increases in serum IL-2 and IFN-γ levels in the severe group were only observed at 4-6 days after disease onset (Figure 3C and 3F). No significant differences in IL4 and TNF-α levels were observed between the two groups during the whole course of the disease (Figure 3D and 3E). All examined cytokines reached similar levels between the severe and mild patient groups at 16 days after disease onset (Figure 3).

**Figure 3.**
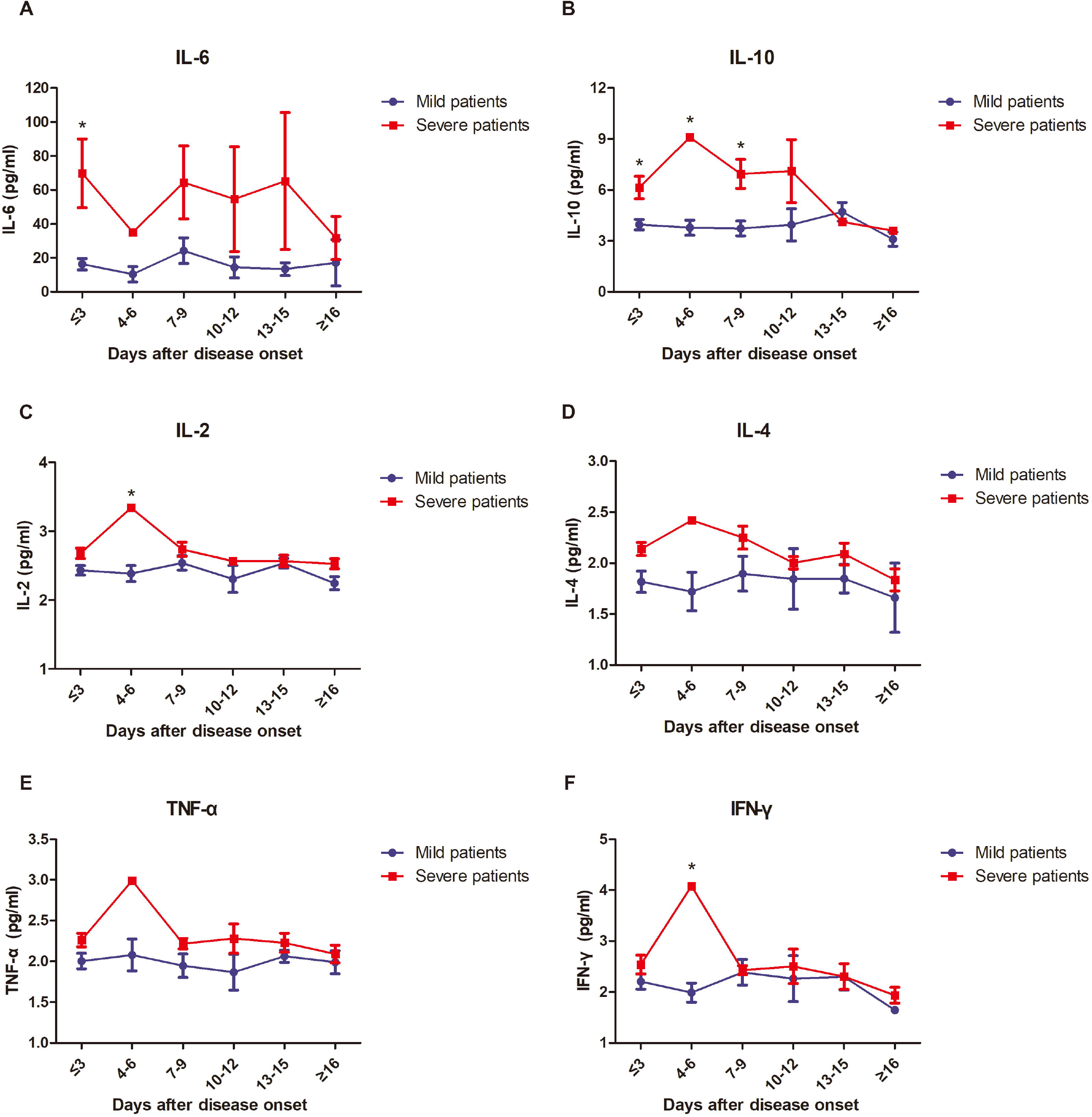
Kinetic analysis of levels of inflammatory cytokines the serum of COVID-19 patients. The concentrations of IL-6 (A), IL-10 (B), IL-2 (C), IL-4 (D), TNF-α (E) and IFN-γ (F) in the serum of mild (blue line) and severe (red line) COVID-19 patients were analyzed at different time points after hospital admission. Error bars, mean ± s.e.m.; *p<0.05.

### Prognostic factors for identification of severe COVID-19 cases

Next, we examined the possibilities of using above-mentioned parameters as prognostic factors for identifying severe cases in COVID-19 patients. PCA was firstly performed by R package “factoextra” to identify correlated variables for distinguishing severe patients from mild patients (Figure 4A). Four mostly contributing variables, neutrophil-to-CD8+ T cell ratio (N8R), neutrophil-to-lymphocyte ratio (NLR), neutrophil counts (NEC) and White Blood Cells counts (WBCC) were selected as potential prognostic factors for further detailed statistical analysis. To assess the diagnostic value of these 4 selected parameters, receiver operating characteristic (ROC) curve and area under ROC curve (AUC) were calculated by R package “pROC” (Figure 4B). The results of this analysis identified N8R with a higher AUC (0.94) than NLR (0.93), NE (0.91) and WBC (0.85). Simultaneously, the cutoff values were calculated from the ROC curves, with a value of 21.9 for N8R (Specificity: 92.6%, Sensitivity: 84.6%), 5.0 for NLR (96.3%, 84.6%), 3.2 for NE (81.5%, 84.6%) and 4.3 for WBC (74.1%, 84.6%) (Figure 4B). The further univariate analysis including the 4 factors of N8R>21.9, NLR>5.0, NE>3.2 and WBC>4.3 were used to calculate odds ratios (ORs) between severe and mild groups. The results were obtained for NLR (OR: 143, 95% Cl: 11.72-1745.3), N8R (OR: 68.75, 95% Cl: 8.55-552.68), NE (OR: 22, 95% Cl: 3.646-132.735) and WBC (OR: 55, 95% Cl: 6.779-446.23) with our patient cohort as predictive factors for severe COVID-19.

**Figure 4.**
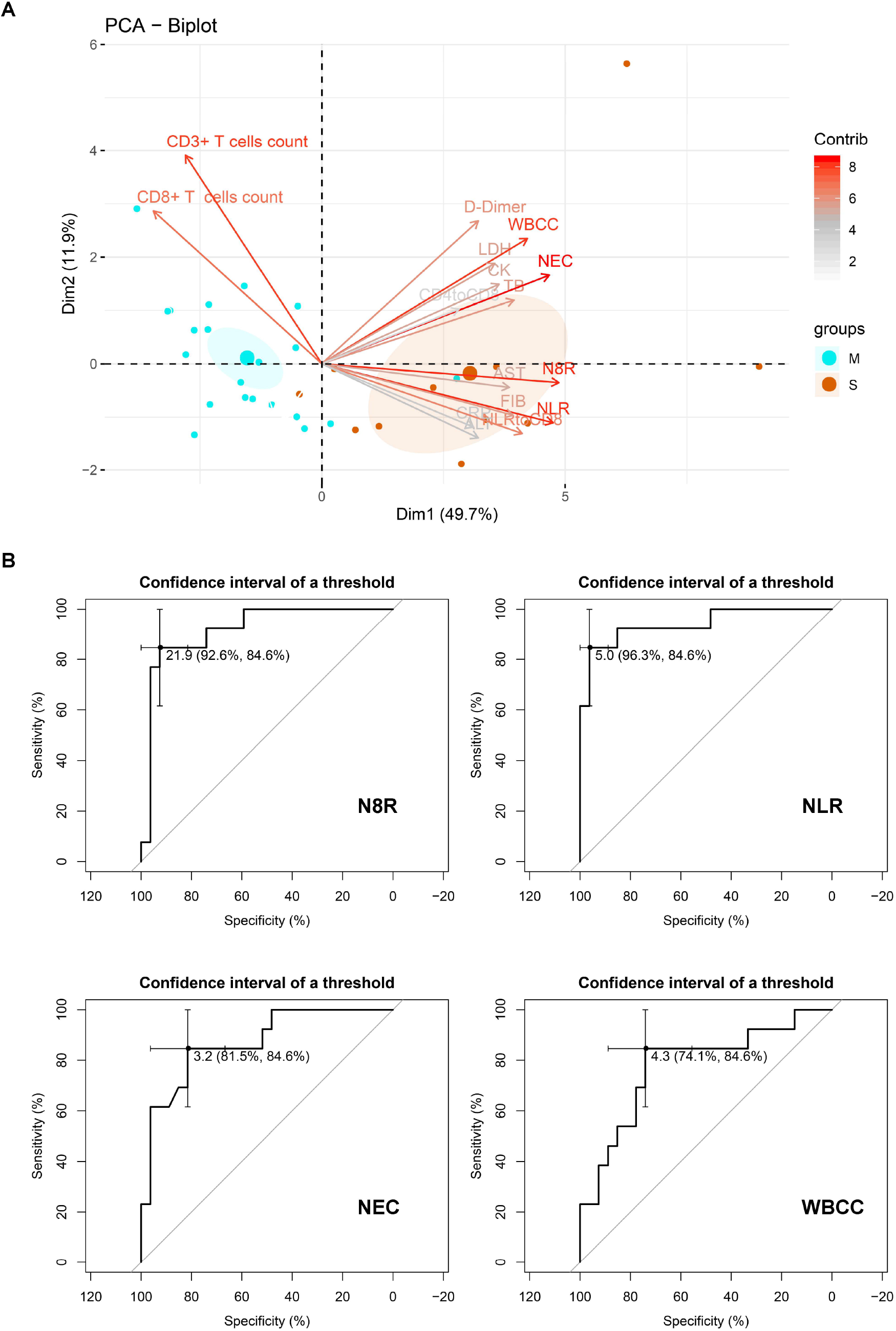
Prognostic factors of severe COVID-19. (A) Principal component analysis was performed by R package “factoextra” to identify correlated variables for distinguishing severe patients from mild COVID-19 patients. Four mostly contributing variables, neutrophil-to-CD8+ T cell ratio (N8R), neutrophil-to-lymphocyte ratio (NLR), neutrophil counts (NE) and White Blood Cells counts (WBC) were identified. (B) ROC curve and AUC were calculated for these 4 selected parameters by using R package “pROC”. The further Logistic regression analysis including the 4 factors of N8R>21.9, NLR>5.0, NE>3.2 and WBC>4.3 was used to calculate OR. The results were obtained for NLR (OR:143, 95% Cl:11.72-1745.3), N8R (OR:68.75, 95% Cl:8.55-552.68), NE (OR:22, 95% Cl: 3.646-132.735) and WBC (OR:55, 95% Cl:6.779-446.23).

## Discussion

In this study, we analyzed the clinical features and immunological characteristics of peripheral blood in patients with COVID-19. Although the majority of the patients did not have an exposure history of the Huanan seafood market in Wuhan, the clinical characteristics of these patients are very similar to those reported in previous studies[2,4,7]. The ages of severe patients are older, and the proportion of underlying diseases is higher, and co-infection also occurs in severe patients. Recent reports show that the lymphocyte counts are normal in COVID-19 patients with mild diseases. In contrast, 63%-70.3% of patients with severe diseases have lymphopenia and the lymphocyte counts in patients with a mortal outcome remain at a low level[4,8]. Our study also confirmed higher rates of developing lymphopenia in severe patients than in mild patients (84.6% vs 44.4%). We found that the development of lymphopenia in severe patients was mainly related to the significantly decreased absolute counts of T cells, especially CD8^+^ T cells, but not to B cells and NK cells. The decrease of T cells in the severe patient group reaches its peak within the first week during the disease course, and then T cell numbers gradually increase from the second week and recover to a comparable level to that of the mild patient group in the third week. All these severe patients included in our study survived the disease, and thus we speculate this course is associated with a favorable outcome in severe COVID-19 patients.

Previous researches on SARS-CoV and MERS-CoV infections have demonstrated the correlation between T cell counts and the severity of the diseases, as well as explored the possible mechanisms[9]. It has been shown that the acute SARS-CoV infection was associated with marked lymphopenia in about 80% of patients, including a dramatic loss of both CD4^+^ and CD8^+^ T cells in comparison with healthy control individuals[10-12]. Decreases in T cell numbers are strongly correlated with the severity of acute phase of SARS disease in patients[11,13]. Lymphopenia is also observed in MERS-CoV infected patients. A detailed clinical study showed that 14 % of MERS patients had leukopenia, while 34 % of the patients had lymphopenia[14]. The mechanism of developing lymphopenia may differ in SARS-CoV and MERS-CoV infections. SARS-CoV cannot productively infect T cells, however, altered antigen presenting cells (APC) function and impaired dendritic cells migration during SARS-CoV infection may result in insufficient T cell priming and thus contribute to decreased numbers of virus-specific T cells in the lungs[15,16]. Moreover, delayed type I interferon response or inflammatory monocyte-macrophages derived pro-inflammatory cytokines could also sensitize T cells to undergo apoptosis[17]. In contrast, MERS-CoV was found to be able to infect many human immune cells, including dendritic cells[18], macrophages[19], and T cells[20]. MERS-CoV infection of T cells results in apoptosis mediated by a combination of extrinsic and intrinsic apoptosis pathways, which is believed to contribute to virus spread and the severe immunopathology[20]. So far, it remains unclear whether SARS-CoV-2 induced T cell contraction is the result of a direct T cell infection or an indirect effect cause by the virus, such as APC function disorder or overactive inflammatory responses. Further studies are needed to investigate the corresponding mechanisms in detail.

Previous studies have shown that elevated levels of proinflammatory cytokines, such as IFN-γ, TNF-a, IL-6 and IL-8, are associated with severe lung injury and adverse outcomes of SARS-CoV or MERS-CoV infection[6,18,19,21]. Our results also demonstrate that severe COVID-19 patients have higher concentrations of IL6, IL10, IL2 and IFN-γ in the serum than mild cases, suggesting that the magnitude of cytokine storm is associated with the disease severity. Additionally, T cells are important for dampening overactive innate immune responses during viral infection[22,23]. Thus, loss of T cells during SARS-CoV-2 infection may result in aggravated inflammatory responses, while restoring T cell numbers may alleviate them. In line with this hypothesis, we observed that the kinetic changes of T cell counts are reversely correlated with the kinetic changes of most examined cytokine levels in the peripheral blood in severe COVID-19 patients. While T cell counts drop to their lowest levels at 4-6 days after disease onset, serum IL-10, IL-2, IL-4, TNF-α and IFN-γ levels reach their peaks. The courses of restoring T cell numbers are associated with decreases of serum IL-6, IL-10, IL-2, IL-4, TNF-α and IFN-γ levels.

Early identification of risk factors for severe COVID-19 patients may facilitate appropriate supportive care and promptly access to the intensive care unit if necessary. A recent study in a 61-patient cohort reported that the NLR was the most useful prognostic factor affecting the prognosis for severe COVID-19[24]. The severity of pathological injury during SARS or MERS correlates with the extensive infiltration of neutrophils in the lung and increased neutrophil numbers in the peripheral blood[17]. Thus, the magnitude of increase in neutrophil counts may suggest the intensity of inflammatory responses in COVID-19 patients. Besides, the magnitude of decrease in lymphocyte counts also indicates the extend of the impairment of immune system by the viral infection. Therefore, NLR may serve as a useful factor to reflect the intensity of imbalance of inflammation and immune responses in COVID-19 patients. In this study, we also screened the potential prognostic factors affecting incidence of severe illness in our patient cohort. Based on our findings with analyzing lymphocyte subsets, we further included the ratio of neutrophils to different lymphocyte subsets as parameters. Our kinetic analysis revealed that CD8^+^ T cells are the major lymphocyte subset which decreases in cell numbers during COVID-19. In line with this finding, our results demonstrate that N8R has even better performance with a higher AUC value than NLR in the ROC curve analysis, and may serve as a more powerful factor than NLR for predicting the severe illness incidence in COVID patients.

In summary, our study of immunological characteristics of the peripheral blood in COVID-19 patients shows that the numbers of neutrophils and T cells, especially CD8^+^ T cells, as well as the levels of inflammatory cytokines in the peripheral blood is dynamically correlated with the severity of the disease. To the best of our knowledge, this is the first work to describe the kinetic changes of lymphocyte subsets and cytokine profiles in COVID-19 patients. Importantly, we identified N8R and NLR as powerful prognostic factors for early identification of severe COVID-19 cases. This work may help to achieve a better understanding of immune function disorder as well as immunopathogenesis during SARS-CoV-2 infection.

## Data Availability

The datasets generated and analyzed during the current study are available from the corresponding author on reasonable request.

## Acknowledgement

We thank all the doctors, nurses, disease control workers, and researchers who have fought bravely and ceaseless against the virus on the frontline during the SARS-CoV-2 epidemic, some of whom lost their lives in doing so. We thank those who have given great and selfless support to the fight against the virus. We thank Ms. Delia Cosgrove and Ms. Ursula Schrammel for language correction of this manuscript.

## Authors contributions

Conceived and designed the experiments: JL, SML, JL, YH, DLY, XZ. Performed the experiments: BYL, HW, WL, QXT, JHY, LZ, LJX, CXG, JT, JZL, JHY, RP, HS, CP, TL, QZ, JW, LX, SHL, BJW, ZHW, CRH, HBZ, RZ, HLZ, XC, PY, BZ, SSH,YWH, SHJ, PW, JAZ, YPL, WXW, LZ, LL, FQZ. Analyzed and interpreted the data: JL, SML, JW. Contributed reagents/materials/analysis tools: XBW, JW, JL, SML. Drafted the manuscript: UD, MJL, JL, DLY, XZ.

